# Incidence of postoperative acute kidney injury is higher in men than women after colorectal surgery – PROSACC: a posthoc analysis of a global, multicenter, randomized controlled trial

**DOI:** 10.1101/2024.09.02.24312832

**Authors:** Robert Frithiof, Mats Enlund, Stephanie Franzén

**Affiliations:** Department of Surgical Sciences, Division of Anesthesiology and Intensive Care, Uppsala University, Uppsala, Sweden; Clinical Research Center, Uppsala University, Västerås, Sweden; Department of Medicine, Division Nephrology, Cardio-Renal Physiology and Medicine, University of Alabama at Birmingham, Birmingham, AL, USA

**Keywords:** acute kidney injury, colorectal, men, women

## Abstract

Postoperative acute kidney injury (AKI) is a common postoperative complication. Approximately 7% of the general, elective, non-cardiac surgical population develop AKI after surgery. The female sex was previously believed to be associated with higher incidence of AKI however more recent literature implies that men have higher risk for AKI. Estrogen has been suggested to have renoprotective properties. We therefore aimed to analyze AKI incidence after colorectal cancer resection surgery in men and women on a global, multicenter, level.

In Sweden and China, a total pf 3255 patients were included in this posthoc analysis based on the CAN-trial (Cancer and Anesthesia: Survival After Radical Surgery - a Comparison Between Propofol or Sevoflurane Anesthesia). Presence of AKI was defined according to KDIGO (Kidney Disease: Improving Global Outcome) criteria for changes in plasma creatinine compared with preoperative values.

After colorectal surgery, 8% of the patients had developed AKI within 10 postoperative days. Within the 4–10-day postoperative timeframe, twice as many men as women (8% vs 4%) had developed AKI and women had a significantly lower likelihood of developing AKI (OR 0.4 [0.2-0.8], p=0.009). In general, the cumulative proportion of developing AKI within 10 days postoperatively was significantly higher in men than women (p=0.037). Moreover, older patients (60+ years) had significantly higher incidence of AKI than those younger than 60 years. This trend was evident in both men and women.

To our knowledge, this is the first ever global, multicenter, randomized controlled trial reporting a sex-difference in AKI incidence after colorectal surgery. Our posthoc analysis reinforces the notion that the male sex is a risk factor for postoperative AKI.

## INTRODUCTION

Postoperative acute kidney injury (AKI) is a common complication after surgery. In the general, elective, non-cardiac surgical population, the incidence of postoperative AKI is approximately 7% 1. The incidence of AKI increase with high-risk surgeries such as major abdominal surgery and cardiovascular surgery ^2–5^. Postoperative AKI also derives with long-term challenges. The odds for chronic kidney disease, end stage renal disease, and in-hospital mortality are significantly increased in patients with AKI compared with those without ^6,7^.

Preoperative independent risk factors for postoperative AKI include obesity, old age, and preexisting comorbidities – such as diabetes and hypertension ^8–10^. Female sex was previously considered a risk factor for postoperative AKI ^11^. However, recent data suggests that the incidence of AKI is actually higher in men than in women. A meta-analysis involving over 6.7 million patients found that hospital-associated AKI was significantly more common in men across various surgical populations, with an overall odds ratio (OR) of 1.23 (1.11-1.36) ^12^. This increased risk was particularly pronounced in non-cardiac surgical populations. Interestingly, men also showed a higher association with AKI in intensive care unit (ICU) populations, whereas this trend was not observed in cardiovascular surgical populations. While not all hospital-associated AKI cases are postoperative, the literature consistently reports a greater association of AKI in men compared to women^13,14^. The underlying mechanisms driving this sex-based difference in AKI susceptibility remain unclear, though sex hormones are thought to play a significant role in the pathogenesis of AKI. Recent conclusions drawn from numerous rodent model studies investigating ischemia-reperfusion-induced AKI support the hypothesis that estrogen exerts a protective effect against AKI, whereas testosterone appears to be detrimental ^15^. However, the precise role of sex hormones in human AKI remains unclear. Coherently, a cross-sectional study in the transgender population revealed that AKI incidence is significantly higher in one of four groups ^16^ – the transfeminine population without hormone therapy ergo the only group at insignificant levels of estrogen. Taken together, large, global, robust, clinical studies are needed to clarify the impact of gender and estrogen levels (postmenopausal women) in the human AKI pathogenesis.

Therefore, we conducted this posthoc analysis on AKI incidence on a global, multicenter, randomized controlled trial with men and women undergoing resection surgery for colorectal cancer. To assess if estrogen is renoprotective in women, age was stratified according to <60 years or postmenopausal (60+ years).

## METHODS

### ETHICS AND STUDY DESIGN

The posthoc analysis was applied on the database from the global, multicenter, randomized controlled trial – Cancer and Anesthesia: Survival After Radical Surgery - a Comparison Between Propofol or Sevoflurane Anesthesia (CAN, NCT01975064) ^17^. The Swedish Ethical Review Authority (Box 2110, 75002, Uppsala, Sweden) reviewed and granted the ethical approval (2021-05316-01) on November 3^rd^, 2021. This posthoc analysis – Propofol and Sevoflurane Anesthesia in Colorectal Cancer Surgery: Incidence of Acute Kidney Injury (PROSACC) – was published on a publicly accessible server before data were accessed (November 22, 2021; Clinical Trials NCT05585866). Inclusion criteria were participant of the CAN trial and colorectal cancer resection surgery. Exclusion criteria was missing preoperative or postoperative plasma creatinine.

### OUTCOMES

Primary outcome was AKI within ten postoperative days. Change in plasma creatinine according to the KDIGO (Kidney Disease: Improving Global Outcome) classification system was used to define AKI. For patients with several creatinine values denoted, the highest value within the timeframe was used for the analysis. Secondary, AKI incidence was stratified by gender (women vs men), and age (<60 vs 60+ years). Outcomes were assessed at different postoperative time points and adjusted for confounding factors.

### DATA MANAGEMENT

The database was extracted from the CAN-study by Uppsala Research Center (retained third party). They performed data cleaning and pseudonymization before providing the database for hypothesis testing.

### STATISTICS

Continuous data are presented as means with standard deviation (SD) and categorical data are presented as total number (%). The Pearson’s Chi^2^ test and Student’s t-test was used for comparing men and women regarding categorical and continuous parameters respectively. Odds ratio analysis for men vs women were performed using the multiple logistic regression analysis with correction for confounding factors. Kaplan Meier diagrams followed by Bonferroni corrected log rank tests were used for analyzing AKI incidence between men vs women and for men vs women with age stratification. StatSoft Statistica (v14.1) was used for all statistical analyses.

## RESULTS

### STUDY DESIGN AND PATIENT CHARACTERISTICS

A total of 3255 patients were included in the PROSACC analysis where 58% (n=1901) were men and 42% (n=1354) were women. Of those 1026 subjects had plasma creatinine values denoted between postoperative day 4-10. A total of 1849 subjects were recruited in 3 different Chinese hospitals and 1406 were recruited in 11 different Swedish hospitals (Table 1). Of all the included subjects, mean age was 65 (12) years, which did not differ between men and women (65 [12] and 66 [13] years, respectively, Table 1). Body mass index (BMI) and weight was significantly higher in men than women (Table 1). Preoperative health status assessed by the American Society of Anesthetists classification system (ASA class) revealed no significant differences between men and women (Table 1). In addition, there were no significant differences between men and women regarding preoperative comorbidities such as hypertension (men: 40%, women: 38%), diabetes (men: 15%, women: 12%), or kidney disease (men: 2%, women: 2%, Table 1).

**Table 1.** Patient characteristics and intraoperative variables. Patient characteristics in all patients and stratified by men and women. Continuous data are presented as mean (SD) and categorical data as number (%) which were analyzed with Student’s t-test and Pearson Chi^2^ test respectively. Significance level was set at 95% and represented as bold red data. BMI – body mass index.

|  | All (n=3255) | Men (n=1901) | Women (n=1354) |
| --- | --- | --- | --- |
| Age, years | 65 (12) | 65 (12) | 66 (13) |
| Weight, kg | 71 (20) | 76 (22) | **63 (13)** |
| BMI, kg/m<sup>2</sup> | 25.0 (8) | 25.4 (10.3) | **24.4 (4.3)** |
| **American Society of Anesthesiologists** |  |  |  |
| Class I | 258 (8%) | 144 (8%) | 114 (8%) |
| Class II | 2535 (78%) | 1469 (77%) | 1066 (78%) |
| Class III | 456 (14%) | 283 (15%) | 173 (13%) |
| Class IV | 5 (0.1%) | 4 (0.2%) | 1 (0.07%) |
| **Center** |  |  |  |
| Shanghai | 1056 | 633 | 423 |
| Xijing | 723 | 411 | 312 |
| Beijing | 70 | 44 | 26 |
| Västerås | 284 | 158 | 126 |
| Uppsala | 16 | 9 | 7 |
| Örebro | 133 | 76 | 57 |
| Linköping | 158 | 96 | 62 |
| Kalmar | 433 | 238 | 195 |
| Skellefteå | 16 | 8 | 8 |
| Sundsvall | 162 | 102 | 60 |
| Helsingborg | 111 | 68 | 43 |
| Västervik | 13 | 9 | 4 |
| Växjö | 51 | 31 | 20 |
| Örnsköldsvik | 29 | 18 | 11 |
| **Comorbidities** |  |  |  |
| Diabetes | 449 (14%) | 292 (15%) | 157 (12%) |
| Hypertension | 1285 (40%) | 759 (40%) | 526 (38%) |
| Kidney disease | 71 (2%) | 45 (2%) | 26 (2%) |

### INTRAOPERATIVE PARAMETERS

Blood loss, fluid balance after surgery, and the length of anesthesia and surgery were all significantly different between women and men. Men lost more blood during surgery than women (+37 [218] mL, p<0.00001, Table 2) and fluid balance after surgery was more positive in men than women (+135 [702] mL, p<0.00001, Table 2). Men also had longer duration of anesthesia (+26 [128] min, p<0.00001) and surgery (+24 [94] min, p<0.00001) than women (Table 2).

**Table 2.** Intraoperative variables. Intraoperative parameters in all patients and stratified by men and women. Continuous data are presented as mean (SD) which were analyzed with Student’s t-test. Simple linear regression was used for correlation R-model. Significance level was set at 95% and represented as bold red data.

|  | All (n=3255) | Men (n=1901) | Women (n=1354) |
| --- | --- | --- | --- |
| **Duration** |  |  |  |
| Surgery, min | 205 (96) | 215 (103) | **191 (85)** |
| Anesthesia, min | 283 (129) | 294 (130) | **267 (127)** |
| **Fluids** |  |  |  |
| Balance, mL | 1327 (712) | 1383 (736) | **1248 (670)** |
| Blood loss, mL | 117 (225) | 132 (244) | **95 (194)** |

### INCIDENCE OF POSTOPERATIVE AKI

Of all included patients, 8% (n=245) developed AKI within 10 postoperative days according to KDIGO classification of plasma creatinine change (preoperative creatinine vs highest postoperative creatinine within 10 days of surgery).

#### Stratifying for gender (men vs women)

Within 10 postoperative days, 6% (n=86) of women and 8% (n=159) of men had postoperative AKI. During postoperative day 1-3, 5% (n=74) of the women had developed AKI and 7% (n=127) of the men. Moreover, at day 4-10, 4% (n=16) of the women and 8% (n=46) of the men had AKI which was a significantly higher incidence amongst men (p=0.013). The cumulative proportion of developing AKI in men and women differs significantly during the 10-day postoperative period (Fig. 1, p=0.037).

**Figure 1.**
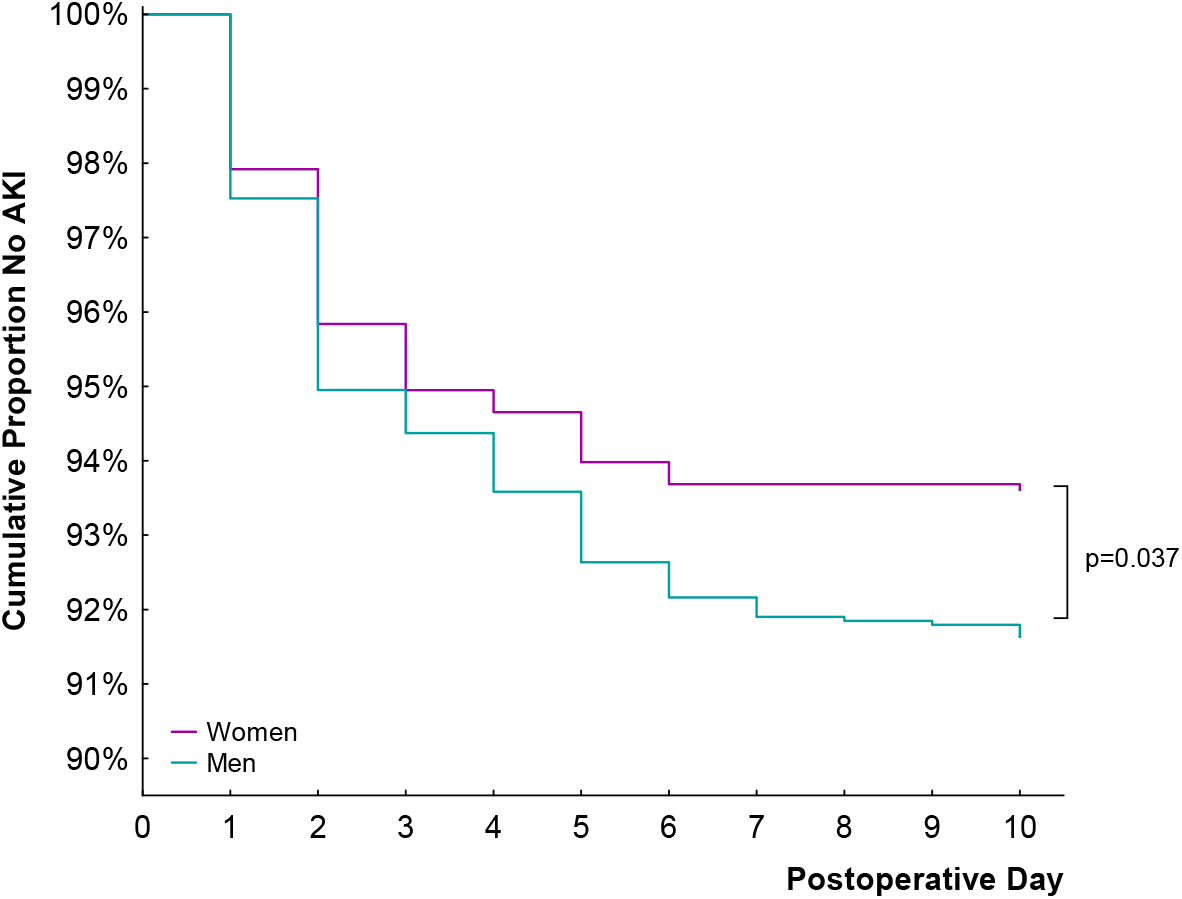
Kaplan Meier graph of cases developing AKI in men (green) and women (purple) over the 10-day postoperative period. Log rank test was performed in order to analyze the curves with a significance level of 95%.

Since AKI incidence was significantly higher in men than women, a multivariate regression analysis was performed with adjustment for potential confounding factors that could affect AKI outcome. There was no difference in AKI incidence between men and women on postoperative days 1-3 (Fig. 2). However, the odds of developing AKI were significantly higher in men compared to women on postoperative days 4-10 (Fig. 2). Specifically, the analysis demonstrated that women had a significantly lower likelihood of developing AKI than men during this period (OR 0.4 [0.2-0.8], p=0.009, Fig. 2), even after adjusting for variables such as higher BMI, greater blood loss, longer surgery duration, and more positive fluid balance.

**Figure 2.**
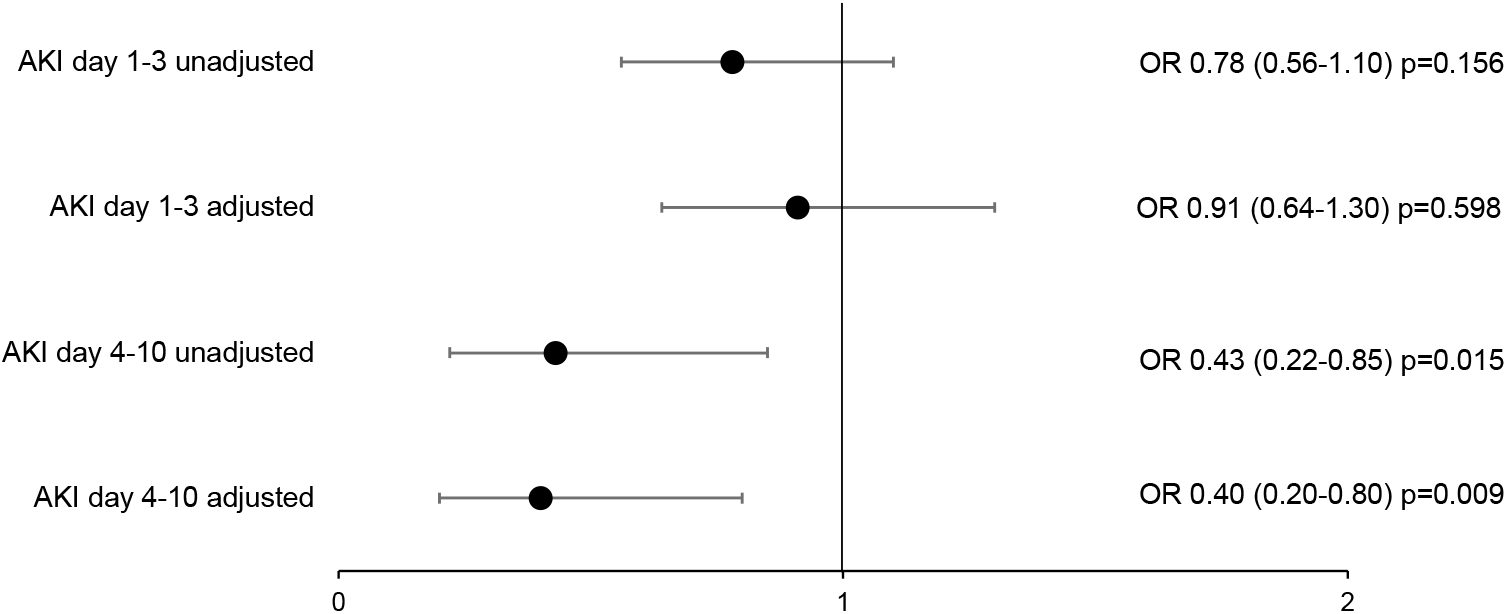
Odds ratio (OR) with 95% confidence interval for women compared with men at postoperative time frame of day 1-3 and 4-10. Adjusted represents values corrected for confounding factors (BMI, blood loss, fluid balance, and anesthesia and surgery duration). Results were derived with the multiple logistic regression analysis with 95% significance level.

#### Stratifying for age (<60 vs 60+ years)

Both men and women were stratified by age into two groups: younger than 60 years and 60 years or older. The cumulative proportion of individuals developing AKI during the 10-day postoperative period differed by age in both sexes (Fig. 3). Among women younger than 60 years, less than 1% (n=3) developed AKI, compared to 9% (n=83) of women aged 60 years or older (p<0.00001). A similar trend was observed in men, with 2% (n=11) of those younger than 60 years developing AKI, compared to 11% (n=148) of men aged 60 years or older (p<0.00001). Although the incidence of AKI was higher in men than in women within both age groups, this difference was not statistically significant.

**Figure 3.**
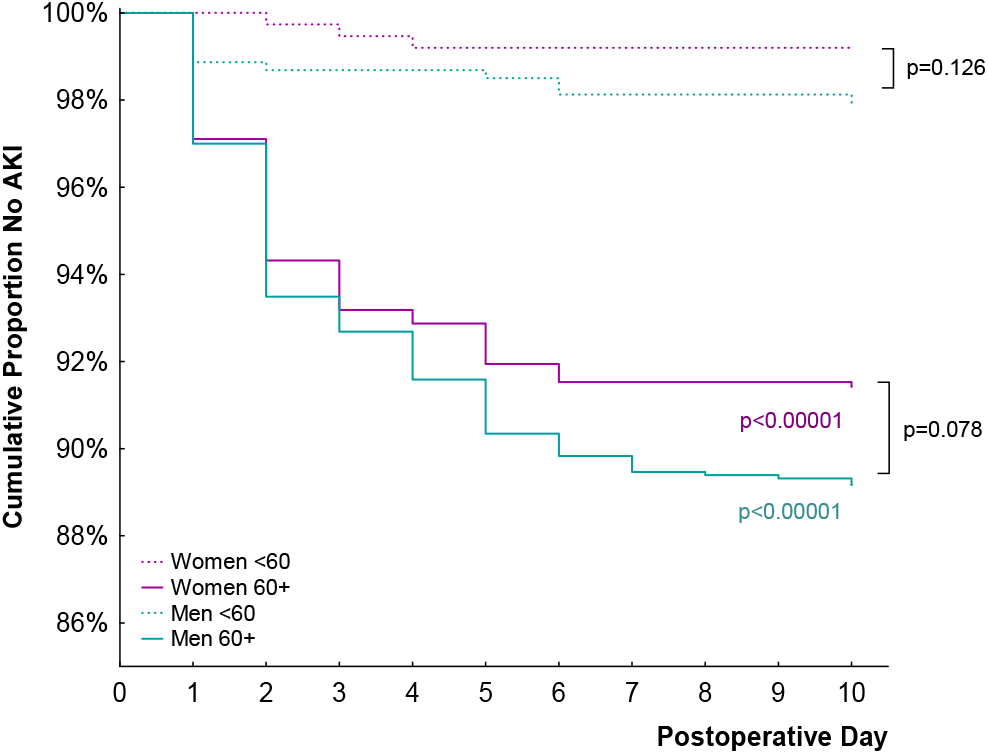
Kaplan Meier graph of cases developing AKI in men <60 years (green dashed), men 60+ years (green), women <60 years (purple dash), and women 60+ years (purple) over the 10-day postoperative period. Log rank tests were performed in order to analyze the curves with a significance level of 98.75% (Bonferroni corrected for multiple comparisons).

## DISCUSSION

This posthoc analysis of Cancer and Anesthesia: Survival After Radical Surgery - a Comparison Between Propofol or Sevoflurane Anesthesia (CAN, NCT01975064) aimed to investigate the difference between men and women regarding AKI outcome after colorectal cancer resection surgery on a global, multicenter level. The main finding was that male sex is associated with a higher incidence of postoperative AKI compared to female sex, with twice as many men as women developing AKI during the 4–10-day postoperative period. Additionally, a significantly higher rate of AKI was observed in older patients compared to younger patients, a trend that was consistent in both men and women. Postoperative AKI remains an under-recognized and under-estimated problem ^9^. The incidence of AKI ranges between 6-18% in the non-cardiac surgical population where major abdominal surgery accounts for the higher percentage ^2,18–21^. Coherently, our analysis aligns with these reports as 8% of the patients developed postoperative AKI. Previously defined risk factors for postoperative AKI include preexisting comorbidities and obesity ^8,10^. In this study, we report that men and women had equal distribution of preexisting comorbidities. However, men had significantly higher BMI which also classified as overweight according to the World Health Organization (≥25 kg/m^2^). According to Chen et al, Chinese men and women average a BMI of 24.9 (3.6) and 23.2 (3.5) respectively ^22^ meanwhile in Sweden, 42% of the women and 63% of the men have an overweight classified BMI ^23^. Both populations indicate that men in general have a higher BMI than women. Moreover, it was shown that a higher BMI is not associated to increased outcome of chronic kidney disease in men but it was in women ^24^. This suggests that regarding kidney disease, overweight is a more significant problem in women than it is in men. Overweight as a risk factor was corrected for in our analysis implying that men have higher risk for AKI independent of BMI.

To our knowledge, this is the first global, multicenter, randomized controlled trial to report a sex dimorphism regarding postoperative AKI after colorectal cancer resection surgery. This is in alignment with a meta-analysis of more than 6.7 million patients worldwide reporting that men have higher association with hospital-associated AKI than women ^12^. Yet the underlying mechanism for sex differences in AKI remains unclear. Mitochondrial function has been suggested as a significant mediator in the pathogenesis of AKI ^25,26^. Furthermore, the literature supports the hypothesis that estrogen has renoprotective properties. It was demonstrated that estrogen via the estrogen-related receptor alpha is a key mechanism in protecting against cisplatin-induced AKI via mitochondrial pathways ^27^. Moreover, a single bolus of exogenous estradiol was tested in mice as part of hemorrhage resuscitation ^28^. Interestingly, this bolus reduced renal injury assessed by lesser fibrosis and reduced renal kidney injury molecule-1 expression. The potentially renoprotective effect of estrogen does not seem to be mediated by the G-protein coupled estrogen receptor 1 ^29^. Another study on mice reported that male mice had more severe injury after ischemia reperfusion (IR) compared with female mice ^30^. Remarkably, castration prior to IR-injury exacerbated the damage in the female mice and administration of exogenous 17beta-estradiol attenuated this injury. This suggests that estrogen, both endogenous and exogenous, can protect the kidney against IR-induced damage. In our analysis, we report that postmenopausal women (60+ years) have higher incidence of AKI than women younger than 60 years. This in in agreement with what was reported after radical nephrectomy ^31^. Additionally, this study showed that postmenopausal women had lower risk of AKI than men in any age category suggesting that other factors than estrogen might play a role in the reported sex dimorphism in AKI. Women younger than 60 years could be pre-, peri- or postmenopausal. Still, a mean age of 57 years was reported for postmenopausal women ^32^, hence 60 years was chosen as age limit to affirm that the majority of the population was lacking estrogen. In our analysis, we discovered an age dependent factor at 60 years that increases the incidence of AKI in both men and women. Which is in accordance with Kim et al. ^31^. The lack of estrogen in the postmenopausal women could be the major mechanism explaining why older women have higher incidence of AKI than younger women. Although, what the aging mechanism for increasing AKI incidence in men remains inconclusive.

### Limitations

The major limitation of this study is that we only use plasma creatinine for defining AKI according to KDIGO criteria. No urine output was denoted in the patient records of the CAN-study. Furthermore, there is a center-specific variation in how many and when postoperative plasma creatinine was analyzed. For some centers, only one postoperative blood sample was taken the first postoperative day – hence any change in plasma creatinine after a few days would not be denoted in the CAN-trial records. Conclusively, this indicates that there could be a larger number of AKI cases than reported. There was a significant difference between men and women regarding perioperative fluid balance, blood loss, and the total duration of anesthesia and surgery. Why this is the case we are not certain. There could be a difference in anatomy between men and women that could result in a longer procedure for men with increased risk of bleeding, hence men will then receive more fluid to account for the blood loss. Even so, the analyses performed are stratified for any perioperative differences and the multiple logistic regression still reveals that men have higher association with AKI. In regard to the limitations, the study also holds strengths. To our knowledge, this is the first global, multicenter, randomized controlled trial to ever report sexual dimorphism regarding postoperative AKI after colorectal surgery.

In conclusion, this post-hoc analysis demonstrates that women have 60% lower odds of developing AKI compared to men during the 4–10-day postoperative period, with twice as many men affected as women. Additionally, women over 60 years of age exhibited a significantly higher incidence of AKI than their younger counterparts, potentially reflecting the loss of estrogen and its renoprotective effects. However, a similar age-related increase in AKI was observed in men, suggesting that the underlying mechanisms remain unclear. This study reinforces the notion that male sex is a significant risk factor for postoperative AKI.

## ACKNOWLEDGEMENTS & CONTRIBUTIONS

We express sincere gratitude to Magnus Ringbom, the main contact person at Uppsala Research Center during the study.

ME provided the database, interpreted data, reviewed the manuscript, and approved the final version. RF assisted in project planning, interpreted data, reviewed the manuscript, and approved the final version. SF developed the hypothesis, planned the project, analyzed and interpreted data, drafted the manuscript, and approved the final version.

## DATA AVAILABILITY STATEMENT

Data cannot be shared openly due to sensitive data regulations.

